# Persisting adaptive immunity to SARS-CoV-2 in Lower Austria

**DOI:** 10.1101/2021.02.18.21251551

**Authors:** Dennis Ladage, Oliver Harzer, Peter Engel, Hannes Winkler, Ralf J. Braun

## Abstract

The prevalence and persistence of adaptive immunity responses following a SARS-CoV-2 infection provides insights into potential population immunity. Adaptive immune responses comprise of antibody-based responses as well as T cell responses mainly addressing viruses and virus-infected human cells, respectively. A comprehensive analysis of both types of adaptive immunity is essential to follow population-based SARS-CoV-2-specific immunity. In this study, we assessed SARS-CoV-2-specific immunoglobulin A (IgA) levels, SARS-CoV-2-specific immunoglobulin G (IgG) levels, and SARS-CoV-2-specific T cell activities in patients who recovered from a COVID-19 infection in spring and autumn 2020. Here we observed a robust and stable SARS-CoV-2-specific adaptive immune response in both groups with persisting IgA and IgG levels as well as stable T cell activity. Moreover, there was a positive correlation of a lasting immune response with the severity of disease. Our data give evidence for a persisting adaptive immune memory, which suggest a continuing immunity for more than six months post infection.

---

The current coronavirus disease 2019 (COVID-19) pandemic is caused by the severe acute respiratory syndrome coronavirus 2 (SARS-CoV-2). Viral infections are combatted by the human immune system, which consists of the rather unspecific innate immune system and the specific adaptive immune system (Sette and Crotty, 2021). Whereas the first system serves as a first line of defense to provide enough time for the induction of adaptive immune responses, adaptive immunity is virus-specific and has the potential to provide long-lasting immunity. Adaptive immunity comprises of the neutralization of viral particles outside cells in combination with the selective removal of virus-infected human cells. B cells produce antibodies that may specifically bind to viral proteins, thereby neutralizing the virus or tagging it for degradation. In contrast, subclasses of T cells may trigger a suicide mechanism in human cells infected by the virus, thereby interfering with viral proliferation.

B cells produce different classes of antibodies at different time points after viral infection. Multimeric immunoglobulin M (IgM) and dimeric immunoglobulin A (IgA) appear early post infection and typically vanish at later stages. In contrast, monomeric immunoglobulin G (IgG) are produced at later stages and are detected for a much longer period. They can be produced by memory B cells when required, thereby contributing to long-lasting immunity. In about 90% of cases individuals infected with SARS-CoV-2 produce antibodies specific for SARS-CoV-2 proteins within the first three weeks post infection. This includes antibodies directed against the SARS-CoV-2 spike protein, which comprises the receptor binding domain (RBD) enabling the virus to access human target cells (Amanat et al., 2020; Long et al., 2020; Premkumar et al., 2020; Zhao et al., 2020). Recent studies demonstrate that SARS-CoV-2-specific dimeric IgA antibodies dominate the early neutralizing antibody response upon infection and are especially potent in SARS-CoV-2 neutralization (Sterlin et al., 2021; Wang et al., 2021). Thus, antibody-based adaptive immunity decisively contributes to deal with SARS-CoV-2 infections, and individuals with enduring high levels of both IgA and IgG antibodies might be better protected than individuals which only show persistent IgG levels. Notably, in our previous studies we observed high levels of SARS-CoV-2-specific IgA antibodies in participants with COVID-19 history, and these antibody class persisted at high levels for more than six months post infection (Ladage et al., 2020a; Ladage et al., 2020b).

T cells are a heterogenous group of immune cells. CD8^+^ T cells, also known as cytotoxic T cells, may specifically trigger cell death in virus-infected cells, thereby preventing viral proliferation. CD4^+^ T cells can differentiate in various cell types with different functions, including the support of cytotoxic CD8^+^T cells and antibody-producing B cells. Both CD4^+^ and CD8^+^T cells can develop into memory cells, thereby critically adding to long-lasting immunity. In SARS-CoV-2 infections the current paradigm suggests that T cell-based immune response precedes antibody-based immune responses (Sette and Crotty, 2021). Moreover, the lack of sufficient T cell-based immune responses is discussed to be causative for severe COVID-19 progression (Sette and Crotty, 2021). In contrast, the rapid SARS-CoV-2 clearance with early induced virus-specific T cells associates with a mild progression of COVID-19 (Tan et al., 2021). Thus, SARS-CoV-2-specific T cell responses are crucial for preventing severe COVID-19 cases with their associated fatal consequences or with detrimental long-lasting health restrictions.

In this study, we analyzed the adaptive immune status of individuals from Lower Austria, which were infected with SARS-CoV-2 in spring 2020 (first epidemic peak) and in autumn 2020 (begin of the second epidemic peak). We included individuals with severe, mild, and asymptomatic disease. We measured the level of SARS-CoV-2-specific IgA and IgG levels and correlated these data with SARS-CoV-2-specific T cell activities. We observed that the adaptive immune response is highly stable, including persistently high levels of IgA, IgG, T cell activities, and that it correlates with the severity of disease progression.

The study was conducted in accordance with the guidelines of the Local Ethics Committee and in approval of the local and national authorities. Blood samples were drawn from peripheral veins by members of the Lower Austrian Red Cross and were analyzed using a semiquantitative enzyme-linked immunosorbent assay (ELISA) kit (Euroimmun AG, Lübeck, Germany), allowing the detection of SARS-CoV-2-specific IgA and IgG antibodies (more precisely, IgA and IgG antibodies specific to the SARS-CoV-2 spike protein, which is necessary for infecting human cells). In addition, SARS-CoV-2-specific T cell activities were determined by incubating isolated T cells with a SARS-CoV-2-specific peptide mix (more precisely, a peptide mix of the SARS-CoV-2 spike protein) and measuring the release of interferon γ by activated T cells using an ELISA system (interferon γ release assay, IGRA) according to the protocol of the manufacturer (SARS-CoV-2-IGRA, Euroimmun AG). The SARS-CoV-2-specific activities of subtypes of both the CD4^+^ and the CD8^+^ T cell populations can be accessed by this method, although the highest response is from CD4^+^ subpopulations (Murugesan et al., 2020; Petrone et al., 2020).

We recruited 152 participants from Lower Austria, comprising of people with previous COVID-19 history, and 13 healthy controls. Among the 152 participants, we collected for 151 participants valid data for both SARS-CoV-2-specific IgA and IgG levels, and for SARS-CoV-2-specific T cell activities. The following results and interpretations refer to these 151 participants.

Diagnosis of a former COVID-19 infection was established by with proof of (former) PCR tests for acute SARS-CoV-2 infection or with positive test for SARS-CoV-2-specific antibodies. Among the 151 participants, 130 (86%) were able to attest their COVID-19 history (Table 1). Among the 130 participants with attested COVID-19 history, 13 (10%) self-declared that they had asymptomatic SARS-CoV-2 infections, whereas 80 (62%) and 31 (24%) self-declared that they suffered from mild and severe COVID-19, respectively. As asymptomatic SARS-CoV-2 infections are expected to be widespread (Petersen et al., 2020), participants with asymptomatic COVID-19 are underrepresented in our test group, whereas those with severe COVID-19 are overrepresented. In 2020, participants with attested COVID-19 history declared having disease symptoms markedly more likely than the control group, like fever (53% vs. 8%), anosmia (42% vs. 0%), ageusia (52% vs. 0%), dyspnea (26% vs. 0%), angina (23% vs. 0%), and weight loss (27% vs. 8%) (Table 2). Thus, the participants with COVID-19 history in our test group experienced typical disease symptoms for COVID-19, as in our previous studies (Ladage et al., 2020a; Ladage et al., 2020b).

**Table 1:**
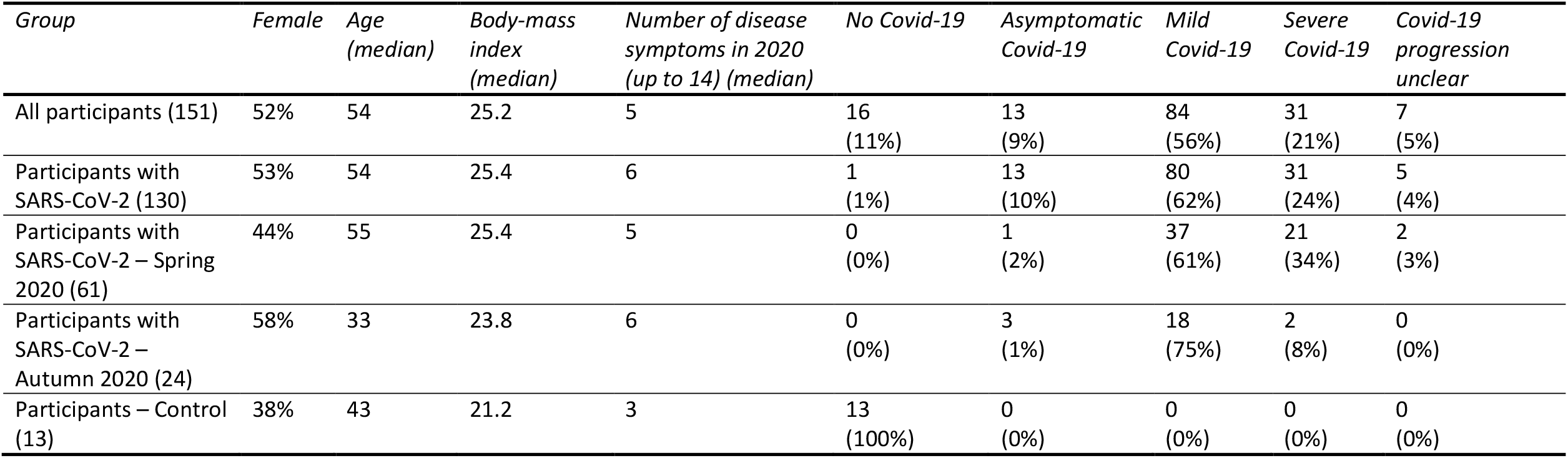
Characteristics of analyzed groups, and severity of disease progression. From “All participants” we have valid data for SARS-CoV-2-specific IgA and IgG levels, and for SARS-CoV-2-specific T cell activities. “Participants with SARS-CoV-2” are defined as a subgroup with experimental evidence of a previous SARS-CoV-2 infection (PCR test, antigen test, or antibody test) in 2020. “Participants with SARS-CoV-2 – Spring 2020” are defined as people with experimental evidence of an acute SARS-CoV-2 infection in Spring 2020 (PCR test in March-May 2020). “Participants with SARS-CoV-2 – Autumn 2020” are defined as people with experimental evidence of an acute SARS-CoV-2 infection in Autumn 2020 (PCR test in September to December 2020). Participants were asked whether they suffered from any of fourteen different disease symptoms in 2020. Data of age, height, weight, disease symptoms in 2020 and severity of Covid-19 disease progression are statements from participants.

| Group | Female | Age<br>(median) | Body-mass<br>index<br>(median) | Number of disease<br>symptoms in 2020<br>(up to 14) (median) | No Covid-19 | Asymptomatic<br>Covid-19 | Mild<br>Covid-19 | Severe<br>Covid-19 | Covid-19<br>progression<br>unclear |
| --- | --- | --- | --- | --- | --- | --- | --- | --- | --- |
| All participants (151) | 52% | 54 | 25.2 | 5 | 16<br>(11%) | 13<br>(9%) | 84<br>(56%) | 31<br>(21%) | 7<br>(5%) |
| Participants with<br>SARS-CoV-2 (130) | 53% | 54 | 25.4 | 6 | 1<br>(1%) | 13<br>(10%) | 80<br>(62%) | 31<br>(24%) | 5<br>(4%) |
| Participants with<br>SARS-CoV-2 – Spring<br>2020 (61) | 44% | 55 | 25.4 | 5 | 0<br>(0%) | 1<br>(2%) | 37<br>(61%) | 21<br>(34%) | 2<br>(3%) |
| Participants with<br>SARS-CoV-2 –<br>Autumn 2020 (24) | 58% | 33 | 23.8 | 6 | 0<br>(0%) | 3<br>(1%) | 18<br>(75%) | 2<br>(8%) | 0<br>(0%) |
| Participants – Control<br>(13) | 38% | 43 | 21.2 | 3 | 13<br>(100%) | 0<br>(0%) | 0<br>(0%) | 0<br>(0%) | 0<br>(0%) |

**Table 2:** Disease symptoms of participants in 2020. Participants were asked whether they suffered from any of fourteen different disease symptoms in 2020. *Asterisk: Disease symptoms, which are markedly more abundant in participants with SARS-CoV-2. Data of disease symptoms are statements from the participants. For details of the analyzed groups and subgroups see legends of Table 1 and Figure 1.

| *Group* | *Fever\** | *Rhinitis* | *Diarrhea* | *Anosmia\** | *Ageusia\** | *Fatigue* | *Dyspnea\** | *Headache* | *Tussis* | *Angina\** | *Sore throat* | *Weight loss\** | *Muscular pain* | *Nausea* |
| --- | --- | --- | --- | --- | --- | --- | --- | --- | --- | --- | --- | --- | --- | --- |
| All participants (151) | 73<br>(48%) | 46<br>(30%) | 43<br>(28%) | 61<br>(40%) | 74<br>(49%) | 104<br>(68%) | 35<br>(23%) | 71<br>(47%) | 77<br>(51%) | 30<br>(20%) | 40<br>(26%) | 36<br>(24%) | 60<br>(39%) | 27<br>(18%) |
| Participants with SARS-CoV-2 (130) | 70<br>(53%) | 37<br>(28%) | 40<br>(31%) | 56<br>(43%) | 68<br>(52%) | 96<br>(73%) | 34<br>(26%) | 62<br>(47%) | 69<br>(53%) | 30<br>(23%) | 36<br>(27%) | 35<br>(27%) | 54<br>(41%) | 23<br>(18%) |
| Participants with SARS-CoV-2 – Spring 2020 (61) | 36<br>(59%) | 13<br>(21%) | 21<br>(34%) | 27<br>(44%) | 34<br>(56%) | 46<br>(75%) | 18<br>(30%) | 28<br>(46%) | 31<br>(51%) | 14<br>(23%) | 14<br>(23%) | 23<br>(38%) | 30<br>(49%) | 11<br>(18%) |
| Participants with SARS-CoV-2 – Autumn 2020 (24) | 9<br>(38%) | 14<br>(58%) | 6<br>(25%) | 10<br>(42%) | 12<br>(50%) | 17<br>(71%) | 4<br>(17%) | 14<br>(58%) | 15<br>(63%) | 2<br>(8%) | 10<br>(42%) | 2<br>(8%) | 8<br>(33%) | 4<br>(17%) |
| Participants – Control (13) | 1<br>(8%) | 5<br>(38%) | 2<br>(15%) | 0<br>(0%) | 0<br>(0%) | 4<br>(31%) | 0<br>(0%) | 5<br>(38%) | 3<br>(23%) | 0<br>(0%) | 3<br>(23%) | 1<br>(8%) | 2<br>(15%) | 3<br>(23%) |

Among the 130 participants with COVID-19 history, the acute COVID-19 could be clearly attributed with PCR tests to spring 2020 (March to May 2020) for 61 (47%) participants, and to autumn (September to mid-December 2020) for 24 (18%) participants (Table 1). Participants with COVID-19 history in autumn 2020 turned out to be markedly younger and less overweighted and suffered in larger part from milder forms of COVID-19 as compared to their counterparts with COVID-19 history in spring 2020 (Table 1, Figure 1).

**Figure 1:**
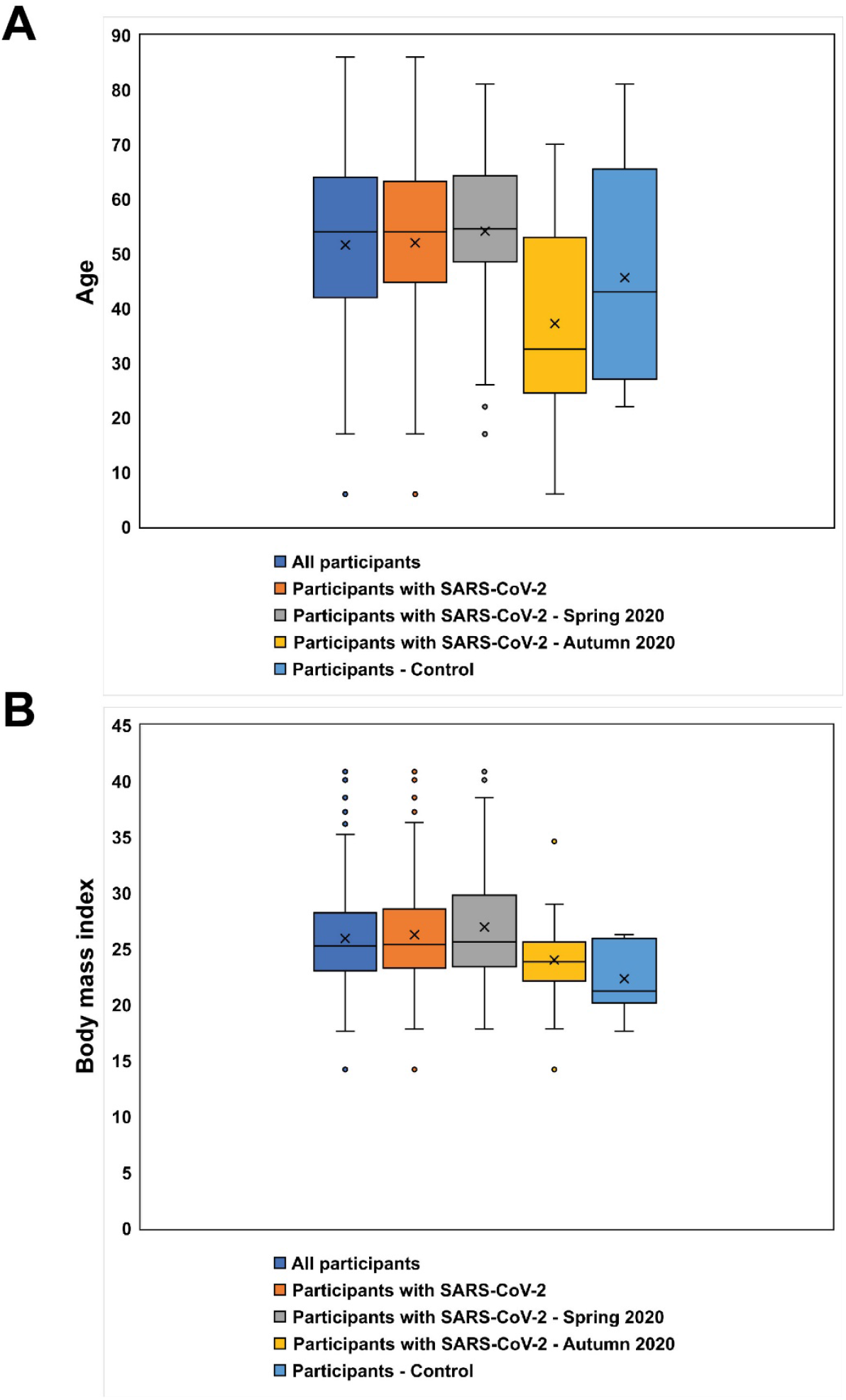
Age and body mass index distribution of groups and subgroups tested for SARS-CoV-2-specific adaptive immunity. Box plots showing (A) age, and (B) body mass index distributions. From “All participants” we have valid data for SARS-CoV-2-specific IgA and IgG levels, and for SARS-CoV-2-specific T cell activities. “Participants with SARS-CoV-2” are defined as a subgroup with experimental evidence of a previous SARS-CoV-2 infection (PCR test, antigen test, or antibody test) in 2020. “Participants with SARS-CoV-2 – Spring 2020” are defined as people with experimental evidence of an acute SARS-CoV-2 infection in Spring 2020 (PCR test in March-May 2020). “Participants with SARS-CoV-2 – Autumn 2020” are defined as people with experimental evidence of an acute SARS-CoV-2 infection in Autumn 2020 (PCR test in September to December 2020). Data of age, height, and weight are statements from participants.

When determining three aspects of SARS-CoV-2-specific adaptive immunity, *i*.*e*., SARS-CoV-2-specific IgA antibody levels, SARS-CoV-2-specific IgG antibody levels and SARS-CoV-2-specific activation of T cells, we observed that 82 of 130 (63%) of participants with COVID-19 history had a robust adaptive immune response comprising all three markers (Figure 2B). Among the 130 participants with COVID-19 history, 121 (86%) had at least two out of three markers, and 126 (97%) had at least one of these markers. Thus, nearly all participants with COVID-19 history showed a SARS-CoV-2 specific adaptive immune response mid-December 2020.

**Figure 2:**
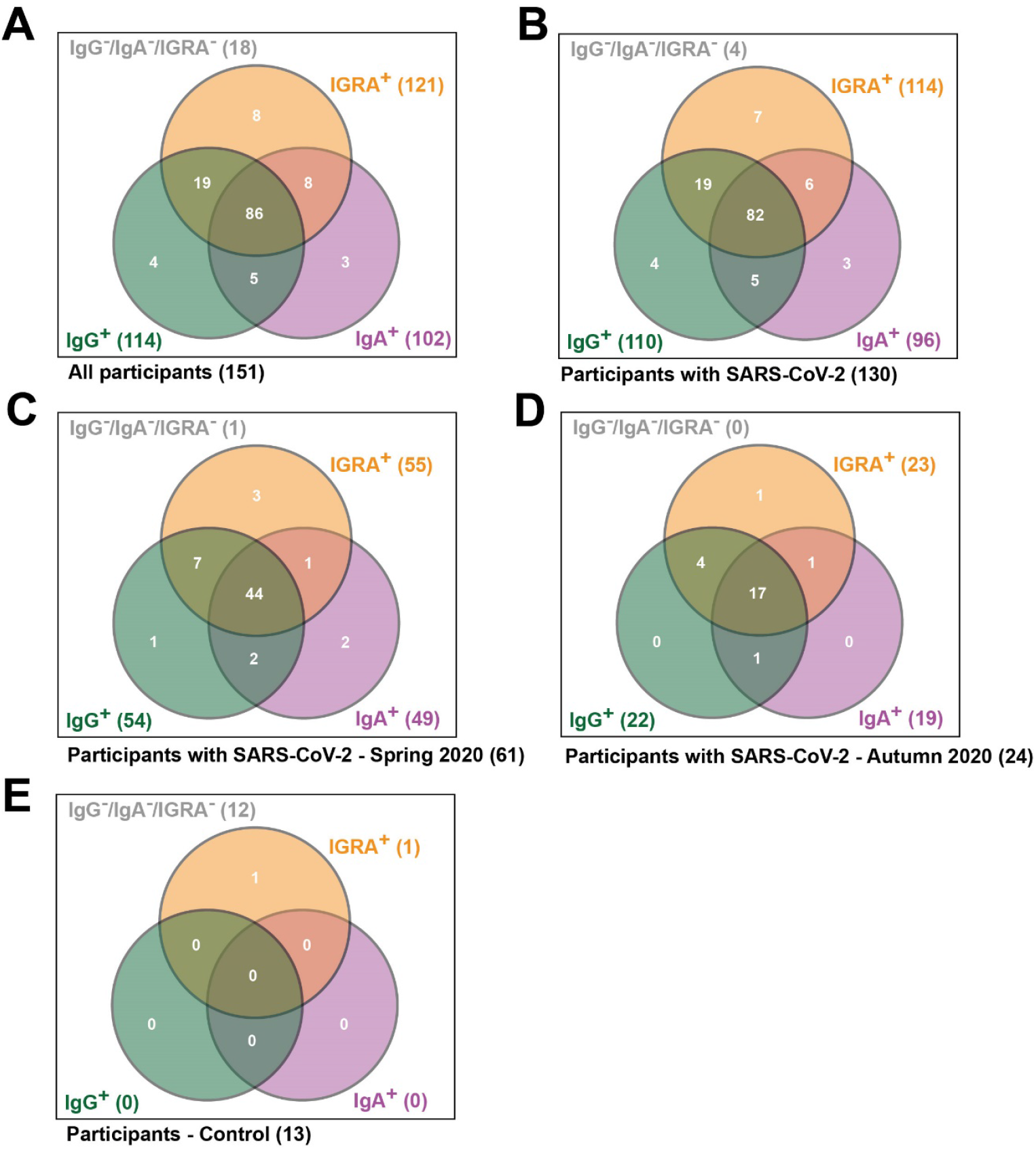
Venn diagrams showing SARS-CoV-2-specific adaptive immune responses. IgA^+^: SARS-CoV-2-specific IgA antibody response. IgG^+^: SARS-CoV-2-specific IgG antibody response; IGRA^+^: SARS-CoV-2-specific T cell response (interferon γ-releasing CD4^+^ and CD8^+^ T cells when triggered with a SARS-CoV-2-specific peptide cocktail). For details of the analyzed groups and subgroups see legends of Table 1 and Figure 1.

When focusing on the participants with acute COVID-19 in spring 2020, we observed that 44 out of 61 participants (72%) had all three markers of SARS-CoV-2-specific adaptive immune response, 54 of 61 (89%) had at least two markers, and 60 out of 61 (98%) had at least one marker (Figure 2B). Notably, these proportions are remarkably similar for participants with acute COVID-19 in autumn 2020 (17 out of 24, 71%, for three markers; 23 of 24, 96%, for at least two markers; 24 of 24, 100%, for at least one marker) (Figure 2C). Thus, the SARS-CoV-2-specific adaptive immune response of participants which suffered from COVID-19 early in 2020 is indistinguishable from the adaptive immune response of those which developed COVID-19 six months later. Our data indicate a very robust long-lasting adaptive immune response with both SARS-CoV-2-specific antibody and T cell activities in participants with COVID-19 history. As the participants with COVID-19 in spring 2020 are older and had an unhealthier form of life (suggested by their higher mean body mass index), both of which may interfere with adaptive immunity (de Heredia et al., 2012; Oh et al., 2019), this stable SARS-CoV-2-specific adaptive immune response is remarkable. On the other hand, participants with COVID-19 in spring 2020 had more severe forms of COVID-19 as compared to participants with COVID-19 in autumn 2020, and this may also influence SARS-CoV-2-specific immunity (Chen et al., 2020).

Consequently, we tested SARS-CoV-2-specific adaptive immunity in participants with severe, mild, and asymptomatic COVID-19 (Figure 3). We observed that 24 out of 31 (77%) participants with severe COVID-19 revealed marked levels of all three measured components of adaptive immunity (SARS-CoV-2-specific IgA, IgG, and T cell activity) (Figure 3A), but only 49 out of 80 (61%) participants with mild COVID-19 (Figure 3B) and only 6 out of 13 (46%) participants with asymptomatic COVID-19 (Figure 3C). Similar effects could be seen when focusing on participants with COVID-19 history in spring 2020 and in autumn 2020 (Figure 3D-G). Thus, the SARS-CoV-2-specific adaptive immune response potentially correlates with COVID-19 severity, with asymptomatic COVID-19 triggering the lowest adaptive immune response. Despite of this finding, many participants with asymptomatic COVID-19 history (9 out of 13, 69%) showed at least two components, and most (11 out of 13, 85%) showed at least one component of SARS-CoV-2-specific adaptive immunity, and (Figure 3C). Thus, SARS-CoV-2-specific adaptive immune response can be seen in nearly all participants with COVID-19 history, but a comprehensive adaptive immune response appears to be more likely in participants with more severe COVID-19.

**Figure 3:**
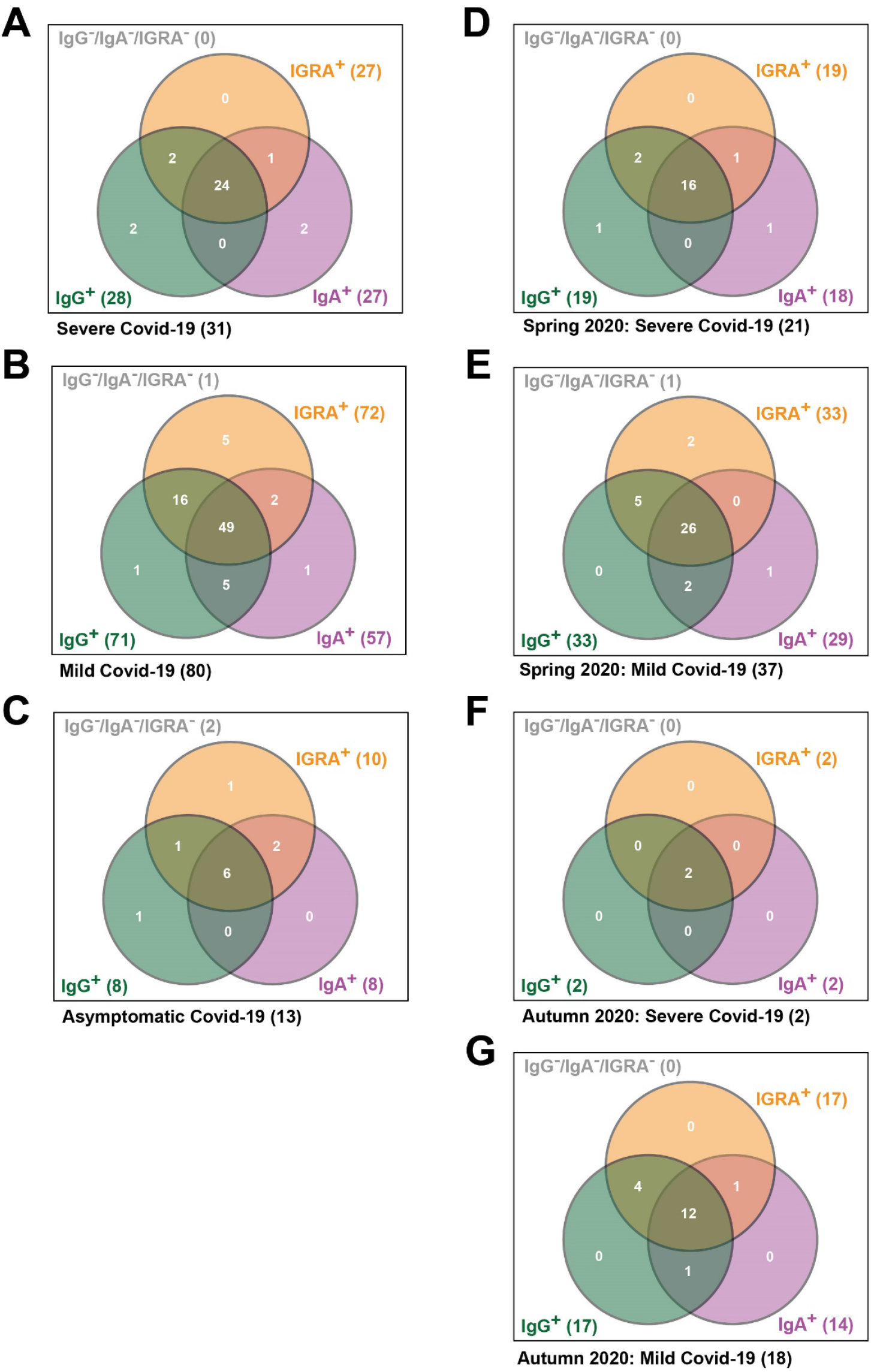
Venn diagrams showing SARS-CoV-2-specific adaptive immune responses in participants with severe, mild and asymptomatic Covid-19 disease progression. (A, B, C): Participants with SARS-CoV-2. (D, E): Participants with SARS-CoV-2 – Spring 2020. (F, G): Participants with SARS-CoV-2 – Autumn 2020. IgA^+^: SARS-CoV-2-specific IgA antibody response. IgG^+^: SARS-CoV-2-specific IgG antibody response; IGRA^+^: SARS-CoV-2-specific T cell response. For details of the analyzed groups and subgroups see legends of Table 1 and Figure 1. Data of Covid-19 disease severity are statements from participants.

In summary, SARS-CoV-2-specific adaptive immune response is still detectable in nearly all participants with a proven COVID-19 history. However, future more comprehensive analysis of SARS-CoV-2-specific adaptive immunity including SARS-CoV-2-specific IgA, IgG, and T cell activities will be of great interest. Especially the SARS-CoV-2-specific immune response after vaccination in comparison to patients with COVID-19 history is a key to understanding whether a similar adaptive immune response is taken place. Alternatively, the antibody-based adaptive immune response might be more prominent upon vaccination at the expense of T cell immunity.

Our previous study (Ladage et al., 2020b) was in line with other studies suggesting that the antibody based immune responses persists for at least several months (Baumgarth et al., 2020; Crawford et al., 2020; Isho et al., 2020; Iyer et al., 2020; Ripperger et al., 2020; Rodda et al., 2020; Wajnberg et al., 2020). Our current findings contribute further evidence that T cell-based immunity markedly contribute to prolonged immune response against SARS-CoV-2 re-infections.

## Data Availability

Data are available from the authors upon request

## Acknowledgements

The authors declare no conflicts of interest.

